# A New Functional F-Statistic for Gene-Based Inference Involving Multiple Phenotypes

**DOI:** 10.1101/2021.05.28.21257997

**Authors:** Adam J. Dugan, David W. Fardo, Dmitri V. Zaykin, Olga A. Vsevolozhskaya

## Abstract

Genetic pleiotropy is the phenomenon where a single gene or genetic variant influences multiple traits. Numerous statistical methods exist for testing for genetic pleiotropy at the variant level, but fewer methods are available for testing genetic pleiotropy at the gene-level. In the current study, we derive an exact alternative to the Shen and Faraway functional F-statistic for functional-on-scalar regression models. Through extensive simulation studies, we show that this exact alternative performs similarly to the Shen and Faraway F-statistic in gene-based, multi-phenotype analyses and both F-statistics perform better than existing methods in small sample, modest effect size situations. We then apply all methods to real-world, neurodegenerative disease data and identify novel associations.

## 1 Introduction

Genetic pleiotropy is the phenomenon where a single gene or genetic variant influences multiple traits [1]. A recent study estimated that more than half of the human genome contains trait-associated loci and that nearly 90% of those loci are shared by more than one trait [2]. A specific example of pleiotropy in neurodegenerative disease is the *MAPT* gene which is a known to contribute risk for several tauopathies and also Parkinson’s disease (which is a condition not linked to tau pathology) [3]. Evidence of pleiotropy has also been found for Alzheimer’s disease (AD) and Parkinson’s disease [4], AD and amyotrophic lateral sclerosis [5], early-onset AD and frontotemporal dementia (FTLD) [6], AD-related psychosis and schizophrenia [7], and limbic-predominant age-related TDP-43 encephalopathy (LATE) and FTLD-TDP [8].

Simulation studies have found that statistical methods designed to test for associations between a single variant and several phenotypes have higher power than single-phenotype analyses, even when only one of the phenotypes is associated with the variant [9, 10]. Examples of single-variant, multi-phenotype methods include multivariate analysis of variance (MANOVA), TATES [11], mv-BIMBAM [12], SCOPA [13], and MultiPhen [14]. Additionally, gene- and region-based methods have also been shown to have improved power over single-variant approaches [15]. Examples of region-based methods include VEGAS [16], GATES [17], SKAT-O [18], and ACAT [19].

Few statistical methods exist for jointly analyzing multiple phenotypes over genomic regions, though some researchers have tried combining single-variant multiple-phenotype methods with multiple-variant single-phenotype methods with some success [20]. In the current study, we develop a novel test statistic for performing multi-phenotype, gene-based tests by leveraging methods from the branch of statistics known as functional data analysis (FDA). We then compare the performance of the novel test statistic to a similar FDA-based test statistic along with two other methods capable of testing for multi-phenotype, gene-based associations. Finally, we apply the methods to investigate potential pleotropic effects for several genes and two related but under-studied neurodegenerative diseases, hippocampal sclerosis (HS) and LATE.

## 2 Methods

### 2.1 Functional F-Statistics

#### 2.1.1 The Function-on-Scalar Regression Model

In order to utilize methods from functional data analysis (FDA), and the function-on-scalar regression (FoSR) model in particular, a reverse regression approach was used where the phenotypes of interest are treated as scalar predictor variables (along with any other adjustors) and the genetic information is treated as a functional outcome. Before creating the functional outcomes, the *g* minor allele counts were first flipped using the approach of Vsevolozhskaya *et al*. to remove spurious noise [21]. Then the flipped minor allele counts were smoothed for each of the *n* individual to create *n* smooth, individual-level genotype functions, *G*_*i*_(*t*). The FoSR model will have the following form:

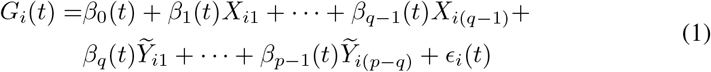

where 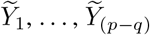 are (*p* − *q*) potentially correlated phenotypes, *X*_1_, …, *X*_(*q*−1)_ are (*q*− 1) adjustment covariates, and *β*_*j*_(*t*) is the association function between the genetic region and the *j*^*th*^ scalar predictor variable. To test the association between the genotype functions, *G*(*t*), and the phenotypes, 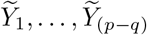, we can compare the full model from Equation 1 to the following reduced model:

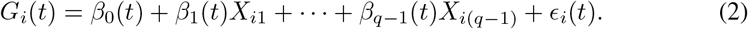

#### 2.1.2 The Shen and Faraway Functional F-Statistic

A functional F-statistic for FoSR models has been proposed by Shen and Faraway [22] with the following form:

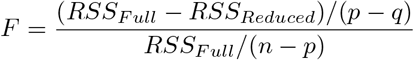

where *n* is the number of observed genotype functions, *p* is the total number of parameters in the full model from Equation 1, *q* is the number of parameters in the reduced model from Equation 2, and *RSS* are the residual sums of squares of a FoSR model and are defined as follows:

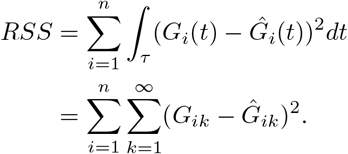

Under the null hypothesis, this F-statistic has the following distribution:

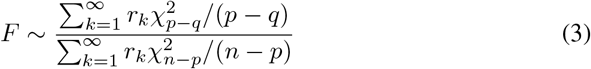

where *r*_*k*_ is the *k*^*th*^ ordered eigenvalue of the variance-covariance matrix of the genotype functions, *G*(*t*). By applying Satterthwaite’s approximation, this distribution can then be approximated by:

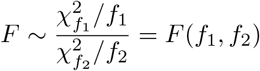

where *f*_1_ = *c*(*p* − *q*), *f*_2_ = *c*(*n* − *p*), and 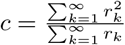

#### 2.1.3 Newly Proposed Functional F-Statistic

Since we expect Satterthwaite’s approximation to be imprecise especially for small p-values which are common in genetic association studies, we proposed an alternative derivation. Instead of using Satterthwaite’s approximation to simplify the distribution of the functional F-statistic from the ratio of two infinite weighted sums of non-independent *χ*^2^ random variables to the ratio of two independent *χ*^2^ random variables, we propose using known properties of the distributions of quadratic forms to evaluate the infinite sums. Specifically, we apply Davies exact method [23, 24] via the `davies()` function from the `CompQuadForm R` package [25] to directly compute the cumulative distribution function (CDF) values of the infinite sums in the numerator and denominator of Equation 3:

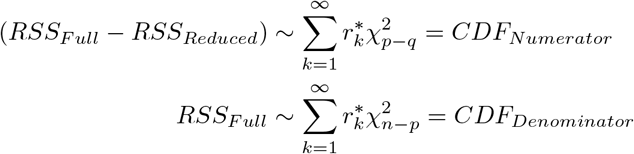

where 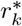 is the *k*^*th*^ ordered eigenvalue of the correlation matrix of the genotype functions, *G*(*t*).

The computed CDF values will be independent and uniformly distributed under the null hypothesis and, thus, can be transformed to follow any distribution. To reflect the traditional F-statistic from multiple linear regression, we transform the numerator and denominator CDF values to 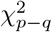 and 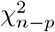 random variables, respectively. Following this approach, the Shen and Faraway functional F-statistic from Equation 3 will have the following distribution under the null hypothesis:

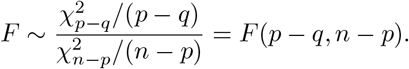

#### 2.1.4 Fitting FoSR Models

Prior to analyzing genomic data with the functional F-statistics, the *g* minor allele counts were first flipped using the approach of Vsevolozhskaya *et al*. to remove spurious noise [21]. Then, generalized additive models were used to fit penalized cubic regression splines to the flipped minor alleles via the `gam()` function from the `gam` and `mgcv R` packages [26, 27]. See Figure 1 for a comparison of raw and flipped smooth genotype functions, *G*_*i*_(*t*), for a region of chromosome 17. Note that the flipped observations no longer necessarily reflect *minor* allele counts, but the underlying associations should be preserved regardless. To reduce the dimensionality of the FoSR models, the individual-level genotype functions, *G*_*i*_(*t*), were then evaluated at *g/*2 equally spaced points and these values were subsequently analyzed. This step isn’t necessary when dealing with small sample sizes or smaller genetic regions as other simulations we conducted (not published here) found it to have little effect on statistical power.

**Figure 1:**
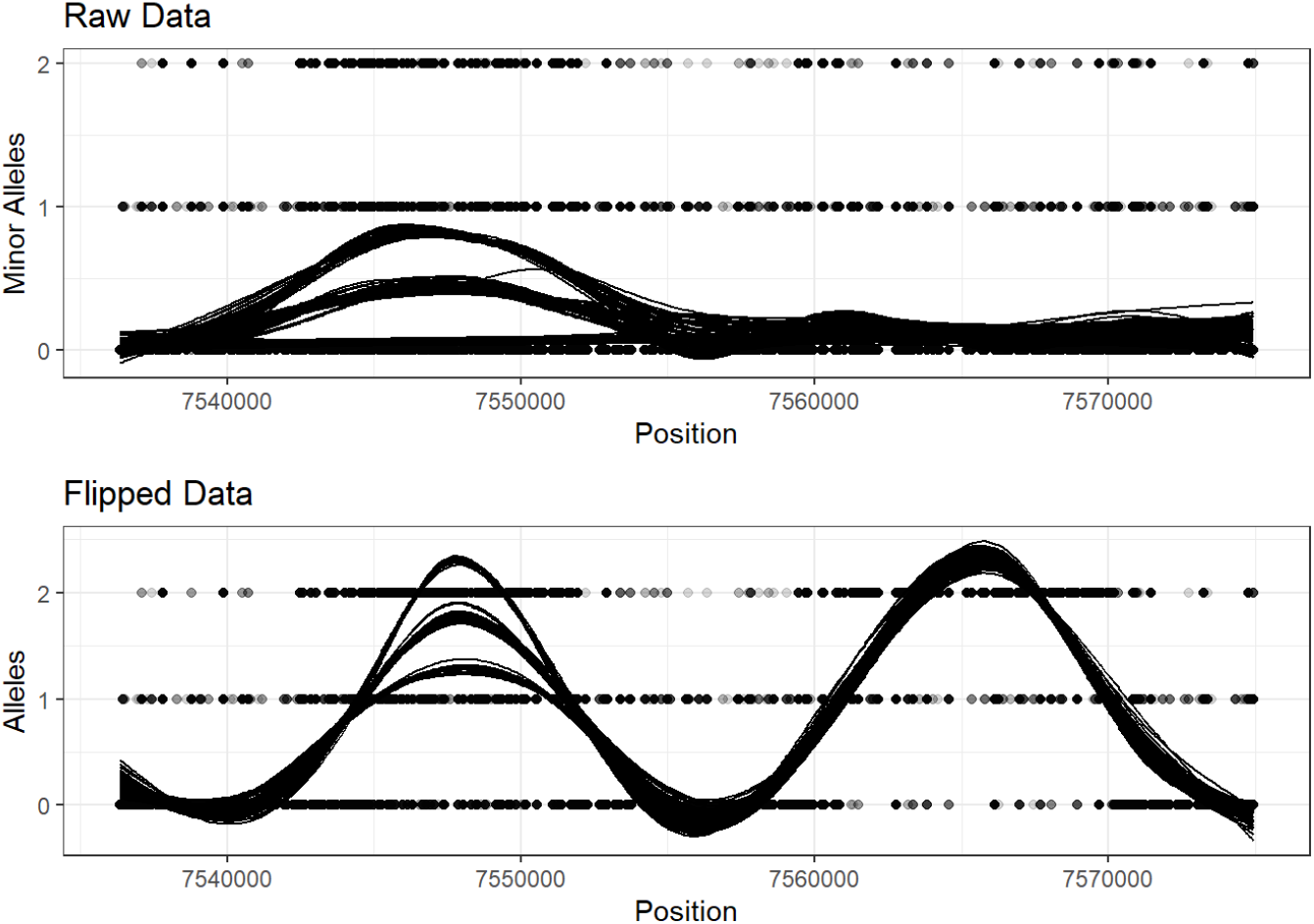
The effect of the minor allele flipping algorithm on the resulting smoothed, individual-level genotype functions, *G*_*i*_(*t*).

FoSR models were fit to the *g/*2 equally spaced *G*(*t*) values using linear mixed effects models via the `nlme R` package [28, 29]. The residual sums of squares were then calculated for both the reduced, intercept-only model and the full model with all phenotypes included as predictors. The empirical variance-covariance matrix of the genotype functions, *G*(*t*), were calculated using the `var.fd()` function from the `fda R` package [30]. Functional F-statistics were then computed as described earlier in Section 2.1.

### 2.2 The Gene Association with Multiple Traits Test

The gene association with multiple traits (GAMuT) test is a statistical method for cross-phenotype analysis using a nonparametric distance-covariance approach that compares similarity in multivariate phenotypes to similarity in genotypes across a gene [31]. Briefly, separate similarity matrices are constructed for the phenotypes and genotypes -***Y*** and ***X***, respectively - either by projection or through the use of kernel functions. Then, each matrix is centered to form ***Y*** _*c*_ and ***X***_*c*_ and the GAMuT test statistic, *T*_*GAMuT*_, is constructed as follows:

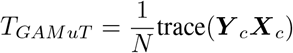

where *N* is the number of individuals included in the analysis. Under the null hypothesis where the two matrices, ***Y*** _*c*_ and ***X***_*c*_, are independent, *T*_*GAMuT*_ follows the same asymptotic distribution as

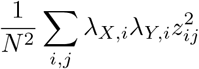

where *λ*_*X,i*_ is the *i*^*th*^ ordered non-zero eigenvalue of ***X***_*c*_, *λ*_*Y,i*_ is the *i*^*th*^ ordered non-zero eigenvalue of ***Y*** _*c*_, and 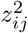 are independent and identically distributed 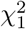 random variables. P-values are then derived using Davies’ exact method [23].

All GAMuT analyses were run in `R` using functions provided by the authors (https://github.com/epstein-software/GAMuT) along with their recommended analytic steps (http://genetics.emory.edu/labs/epstein/software/gamut/GAMuT-example-analysis.html). Phenotype similarity matrices, ***Y***, were constructed such that ***Y*** = ***P*** (***P*** ^*T*^ *P*)^−1^***P*** ^*T*^, where ***P*** is a matrix of phentypes. The raw (i.e., un-flipped and un-smoothed) minor allele counts were used in these analyses. In the applied analyses, where some genotype values were missing for some individuals, variants with any missing values were excluded from the GAMuT analyses.

### 2.3 The Aggregated Cauchy Association Test

The aggregated Cauchy association test (ACAT) is a method that works by converting SNP-level p-values into Cauchy-distributed random variables [19]. Since the sum of dependent Cauchy random variables is identical to the sum of independent Cauchy random variables, no additional information on the linkage disequilibrium or correlation of the SNPs is needed making the method extremely fast [32]. While ACAT was originally developed for the purposes of testing rare variant associations, the authors state that the method can also be applied to common variants.

The ACAT test statistic, *T*_*ACAT*_, has the following form:

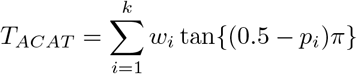

where *k* is the number of SNP-level p-values, *p*_*i*_ is the *i*^*th*^ SNP-level p-value, and *w*_*i*_ is the non-negative weight for the *i*^*th*^ p-value. Note that tan (0.5 − *p*_*i*_)*π* will be Cauchy distributed if *p*_*i*_ is from the null distribution [32]. Then, based on the cumulative density function of the Cauchy distribution, the overall ACAT p-value can be approximated by

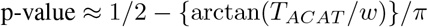

where 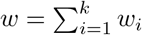 [19].

In our analyses, we first fit regression models to each SNP and phenotype using linear regression models assuming an additive mode of inheritance via the `lm()` function in `R`. The raw (i.e., un-flipped and un-smoothed) minor allele counts were used. Then, we applied ACAT with equal weights to combine SNP-level p-values into a single gene-level p-value for each phenotype. The phenotype-level ACAT p-values were then combined via the minimum-p method to obtain a single, multi-phenotype, gene-level p-value. In the context of Equation 1, the smallest gene-level, phenotype-specific p-value would be multiplied by the number of phenotypes analyzed, (*p*−*q*), to obtain the final p-value. While this approach is expected to be conservative since it fails to take into account the correlation that may exist among phenotypes, it is computationally efficient and provides an analytic baseline for comparing the performance of other statistical methods. All ACAT analyses were run in `R` using functions provided by the authors (https://github.com/yaowuliu/ACAT).

### 2.4 Simulations

#### 2.4.1 Data Simulation

Similar to other simulation studies [33], we utilized realistic linkage disequilibrium patterns by using data from the 1000 Genomes Project [34] for a 100 Kb region of chromosome 17 which included 12,735 SNPs spanning from the *FGF11* gene to the *NDEL1* gene. The selection of this region of the genome was arbitrary, but we expect the linkage disequilibrium structure of this region to be representative of, and generalizable to, other regions of the genome.

For each simulation, we randomly selected a window of width *m* Kb (where *m* ∈ {10, 25, 50}) containing *g* consecutive SNPs for *n* individuals sampled with replacement from the 1,092 available individuals (where *n* ∈ {100, 250, 500}). The resulting matrix was defined to be ***G***_*n*×*g*_ and standard quality control procedures were then applied such as removing SNPs with minor allele frequencies less than 0.01 and correlations with other SNPs greater than 0.99.

Within each window, we then selected *c* SNPs (where *c* ∈ {1, 5, 10}) to be causally associated with all five phenotypes. Causal effects, ***β***_*g* ×5_, were simulated for all SNPs from a normal distribution with *µ* = 0 and

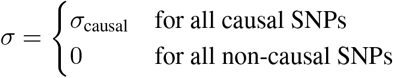

where *σ*_causal_ ∈ {0.05, 0.10, 0.15, 0.25, 0.50, 1.00}. Continuous phenotypes, 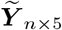, were then generated using the following model:

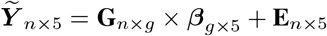

where **E**_*n*×5_ is a matrix of errors that follows a multivariate normal distribution with ***µ*** = 0 and **σ** = *σ*_error_***R***, *σ*_error_ = 1, and ***R*** is a 5 × 5 matrix of the correlations among the five continuous phenotypes. Multivariate normal observations were generated using the `mvrnorm()` function from the `MASS R` package [35].

Phenotypes were simulated assuming an underlying equicorrelation structure where all off-diagonal values of ***R*** were equal to *ρ*, where *ρ* ∈ {0, 0.5}. Disturbance was also added to the underlying correlation matrix, ***R***, for some simulations so that the methods could be systematically tested in situations with unstructured correlation structures. To create a disturbed correlation matrix, a vector was created consisting of five random values from a continuous Uniform(—*b, b*) distribution, where *b* ∈ {0, 5}. Then, this vector was multiplied by its transpose to create a 5 ×5 square matrix, ***B***, of rank 1. Finally, ***B*** was added to ***R*** and the resulting matrix was converted to a correlation matrix to create a disturbed underlying correlation matrix, ***R***^*^, for simulating phenotypes.

For Type-I error simulations where *σ*_causal_ = 0 for all SNPs, a total of 10,000 simulations were run for each scenario. For all power simulations, a total of 1,000 simulations were run for each scenario.

The performance of the functional F-statistics were compared to two other multi-phenotype, gene-based methods: ACAT and GAMuT. For all of the comparison methods, the raw (i.e., not flipped and not smoothed) genetic data were used. SNP-level analyses for ACAT were performed separately for each phenotype assuming an additive mode of inheritance using linear regression models via the `lm()` function in `R`.

## 3 Results

### 3.1 Simulations

#### 3.1.1 Type I Error

All methods had relatively conservative Type I error rates across the simulation scenarios, though they all tended to become more accurate as the sample size, *n*, and gene size, *m*, increased. See Table 1 for Type-I error rates at *α* = 0.05 for each method stratified by gene size, number of observations, correlation among phenotypes, and correlation disturbance.

**Table 1.** Type 1 error at *α* = 0.05 for each method stratified by gene size, number of observations, correlation among phenotypes, and correlation disturbance.

| Gene Size | No. of Obs. | Phenotype Corr. | Corr. Disturbance | ACAT | GAMuT | Shen-Faraway F | New F |
| --- | --- | --- | --- | --- | --- | --- | --- |
| 10,000 | 100 | 0.0 | 0 | 0.052 | 0.034 | 0.030 | 0.022 |
| 10,000 | 100 | 0.0 | 5 | 0.034 | 0.038 | 0.028 | 0.019 |
| 10,000 | 100 | 0.5 | 0 | 0.048 | 0.035 | 0.031 | 0.021 |
| 10,000 | 100 | 0.5 | 5 | 0.037 | 0.034 | 0.032 | 0.024 |
| 10,000 | 250 | 0.0 | 0 | 0.049 | 0.047 | 0.038 | 0.030 |
| 10,000 | 250 | 0.0 | 5 | 0.040 | 0.041 | 0.039 | 0.031 |
| 10,000 | 250 | 0.5 | 0 | 0.050 | 0.042 | 0.036 | 0.027 |
| 10,000 | 250 | 0.5 | 5 | 0.037 | 0.043 | 0.040 | 0.030 |
| 10,000 | 500 | 0.0 | 0 | 0.054 | 0.049 | 0.044 | 0.036 |
| 10,000 | 500 | 0.0 | 5 | 0.037 | 0.046 | 0.040 | 0.034 |
| 10,000 | 500 | 0.5 | 0 | 0.051 | 0.046 | 0.040 | 0.034 |
| 10,000 | 500 | 0.5 | 5 | 0.035 | 0.050 | 0.041 | 0.035 |
| 25,000 | 100 | 0.0 | 0 | 0.051 | 0.032 | 0.045 | 0.035 |
| 25,000 | 100 | 0.0 | 5 | 0.041 | 0.027 | 0.045 | 0.038 |
| 25,000 | 100 | 0.5 | 0 | 0.047 | 0.028 | 0.040 | 0.035 |
| 25,000 | 100 | 0.5 | 5 | 0.037 | 0.029 | 0.045 | 0.038 |
| 25,000 | 250 | 0.0 | 0 | 0.053 | 0.041 | 0.044 | 0.037 |
| 25,000 | 250 | 0.0 | 5 | 0.041 | 0.043 | 0.047 | 0.040 |
| 25,000 | 250 | 0.5 | 0 | 0.050 | 0.039 | 0.049 | 0.043 |
| 25,000 | 250 | 0.5 | 5 | 0.037 | 0.040 | 0.044 | 0.039 |
| 25,000 | 500 | 0.0 | 0 | 0.053 | 0.045 | 0.048 | 0.044 |
| 25,000 | 500 | 0.0 | 5 | 0.039 | 0.045 | 0.054 | 0.051 |
| 25,000 | 500 | 0.5 | 0 | 0.050 | 0.044 | 0.048 | 0.042 |
| 25,000 | 500 | 0.5 | 5 | 0.034 | 0.045 | 0.048 | 0.045 |
| 50,000 | 100 | 0.0 | 0 | 0.054 | 0.023 | 0.049 | 0.042 |
| 50,000 | 100 | 0.0 | 5 | 0.038 | 0.022 | 0.047 | 0.041 |
| 50,000 | 100 | 0.5 | 0 | 0.048 | 0.026 | 0.047 | 0.039 |
| 50,000 | 100 | 0.5 | 5 | 0.036 | 0.026 | 0.050 | 0.043 |
| 50,000 | 250 | 0.0 | 0 | 0.053 | 0.037 | 0.050 | 0.046 |
| 50,000 | 250 | 0.0 | 5 | 0.037 | 0.034 | 0.053 | 0.048 |
| 50,000 | 250 | 0.5 | 0 | 0.051 | 0.035 | 0.051 | 0.045 |
| 50,000 | 250 | 0.5 | 5 | 0.037 | 0.035 | 0.049 | 0.045 |
| 50,000 | 500 | 0.0 | 0 | 0.049 | 0.042 | 0.050 | 0.048 |
| 50,000 | 500 | 0.0 | 5 | 0.037 | 0.038 | 0.048 | 0.042 |
| 50,000 | 500 | 0.5 | 0 | 0.048 | 0.041 | 0.050 | 0.045 |
| 50,000 | 500 | 0.5 | 5 | 0.040 | 0.040 | 0.052 | 0.046 |

Under simulation scenarios with no correlation disturbance, ACAT tended to have the best control of Type I error especially for smaller sample sizes. However, as the sample size increased and as disturbance was added to the correlation, the functional F-statistics tended to perform better. Interestingly, whenever disturbance was added, ACAT’s Type I error tended to suffer while GAMuT and the functional F-statistics tended to be either unaffected or become more accurate. Notably, the Type I error rate of the newly derived functional F-statistic was consistently lower than that of the Shen and Faraway functional F-statistic which made it more conservative in all but one scenario (with a gene size of 25,000 bases, sample size of 500, no correlation among phenotypes, and correlation disturbance).

#### 3.1.2 Power

No single method had the highest power across all simulation scenarios, but some general trends were apparent. Overall, power increased for all methods as the sample size, gene size, number of causal variants, and causal effect size increased. Additionally, all methods tended to achieve their lowest power in scenarios where the phenotypes were independent (*ρ* = 0, and *b* = 0) while they tended to achieve their highest power in scenarios with high amounts of correlation (*ρ* = 0.5, and *b* = 5). Differences between the methods were observed based on causal effect sizes and correlation structures.

For modest causal effect sizes (*σ*_causal_ *<* 0.25), the functional F-statistics were the most powerful methods regardless of gene size, number of causal variants, and correlation structure (Figure 2). The performance of the functional F-statistics were similar to one another, though the Shen and Faraway F-statistic tended to have slightly higher power for the smallest causal effect sizes. While ACAT and GAMuT performed worse than the functional F-statistics for modest effect sizes, especially at smaller sample sizes, their performance relative to one another differed substantially based on the underlying correlation structure. ACAT was far more powerful than GAMuT when the phenotypes were independent (*ρ* = 0, and *b* = 0) and GAMuT was far more powerful than ACAT when there were high amounts of correlation among the phenotypes (*ρ* = 0.5, and *b* = 5). Under their respective most powerful correlation structures, ACAT and GAMuT were able to match the performance of the functional F-statistics at the largest sample size (*N* = 500).

**Figure 2:**
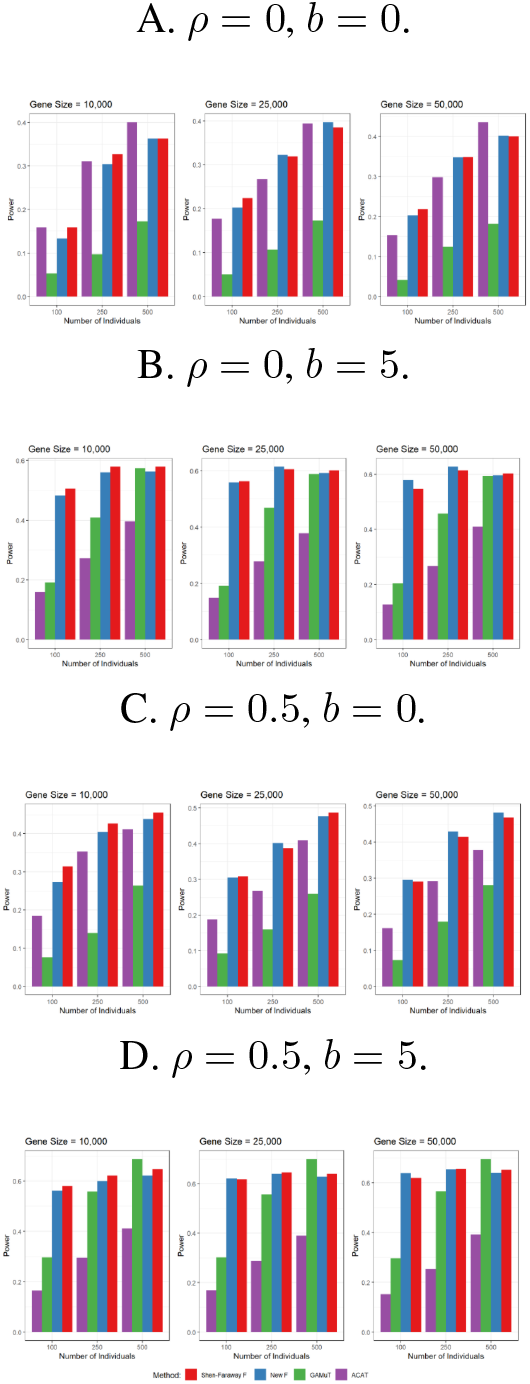
Statistical power for simulations assuming 5 continuous phenotypes, 5 causal variants, and causal effect sizes simulated from a normal distribution with *µ* = 0 and *σ*_causal_ = 0.15.

As the the causal effects increased (*σ*_causal_ ≥0.25), ACAT and GAMuT began outperforming the functional F-statistics especially at larger sample sizes (Figure 3). Similar to the modest effect size scenarios, ACAT tended to be more powerful than GAMuT when the phenotypes were independent (*ρ* = 0, and *b* = 0) and GAMuT tended to be more powerful than ACAT when there were high amounts of correlation among the phenotypes (*ρ* = 0.5, and *b* = 5). Notably, even with stronger causal effects, the functional F-statistics still tended to be perform similar to, or sometimes even better than, the most powerful method at the smallest sample size. Furthermore, while the relative performances of ACAT and GAMuT varied based on the underlying correlation structure, the functional F-statistics were more stable and tended to be the second and third most powerful methods at larger sample sizes.

**Figure 3:**
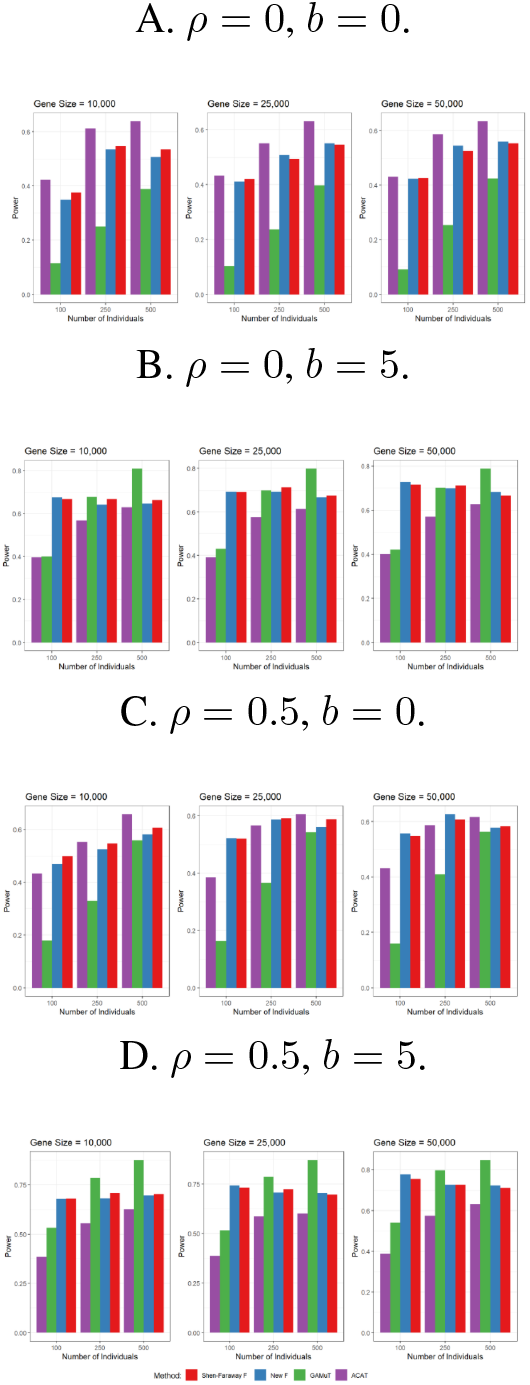
Statistical power for simulations assuming 5 continuous phenotypes, 5 causal variants, and causal effect sizes simulated from a normal distribution with *µ* = 0 and *σ*_causal_ = 0.25.

### 3.2 Application to Neurodegenerative Disease Data

Several genes have been implicated for hippocampal sclerosis (HS), a neurodegenerative disease characterized by severe neuronal cell loss and gliosis in the hippocampus, including: *KCNMB2, TMEM106B, ABCC9 GRN*, and *APOE*. Our understanding of HS has evolved in recent years and what was once considered “HS” is now understood to include several distinct yet related neuropathological diseases, including limbic-predominant age-related TDP-43 encephalopathy (LATE). While genetics are known to play a role in the development of neurodegenerative diseases, the autopsy-based data necessary to definitively diagnose neuropathological changes associated with these conditions is scarce. Therefore, to better understand the genetics underlying the original HS risk genes, we tested for associations between these genes and the more specific autopsy-derived, HS-related neuropathological endophenotypes. If these associations fail to replicate, then we would be inclined to conclude that the genes are associated with some other aspect of HS outside of its neuropathological presentation.

Phenotypic data from the National Alzheimer’s Coordinating Center (NACC) were linked with genotype data from the Alzheimer’s Disease Genetics Consortium imputed using the Haplotype Reference Consortium (ADGC-HRC) [36, 37, 38]. Individuals who died at age 65 years or older were included in this analysis. Similar to other studies of NACC participants [39], individuals were excluded if at least one of 19 rare brain diseases were diagnosed or if they were missing any adjustment variables or all of the neuropathological endophenotypes being analyzed. Each gene was defined based on the canonical transcripts using the hg19 gene range list from PLINK (https://www.cog-genomics.org/plink/1.9/resources) and was flanked by an additional 10kb on both ends to capture potential regulatory variants. ACAT, GAMuT, the Shen-Faraway functional F-statistic, and our newly derived functional F-statistic were applied to the data to test for joint associations between HS and LATE and each gene. Statistical significance was defined to be *p <* 0.05. Note that variants with any missing values were excluded from the GAMuT analyses since it requires the genetic data to have no missingness.

Several genes were found to have a joint association with both HS and LATE, including *TMEM106B, GRN*, and *APOE*. The complete results may be found in Table 2. GAMuT failed to detect any gene-based associations, while both functional F-statistic methods and ACAT were able to detect gene-based associations of HS and LATE with the *TMEM106B* and *APOE* genes. Notably, the only method to detect a gene-based association between HS and LATE and *GRN* was the newly derived functional F-statistic.

**Table 2.**
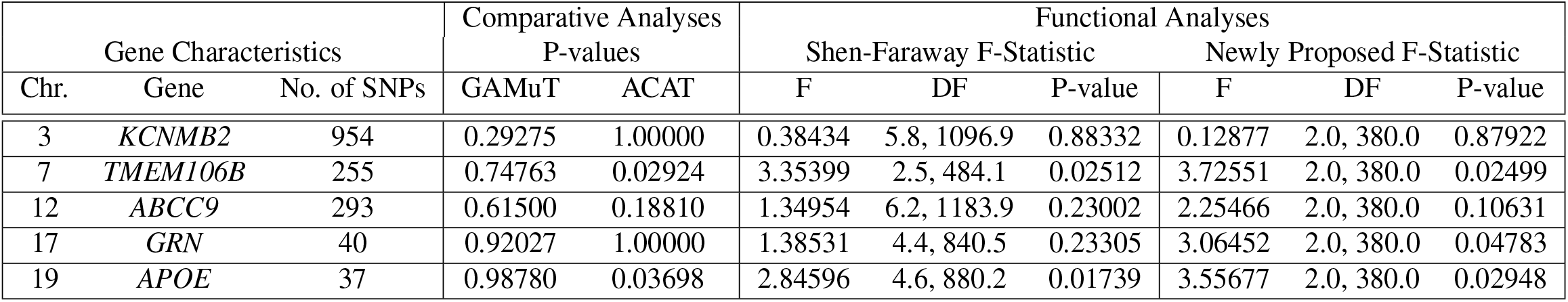
Gene-based results from the applied analysis looking at the pleiotropic effects of several genes on HS and LATE.

## 4 Discussion

By leveraging known properties of quadratic forms, we were able to develop an alternative derivation for the distribution of the functional F-statistic for FoSR models. Through simulation studies, we showed that this functional F-statistic performs similarly to the existing Shen and Faraway F-statistic and both F-statistics can outperform other statistical methods designed for testing gene-level pleiotropy. While ACAT and GAMuT tended to be more powerful in situations with stronger causal effects, the functional F-statistics performed better in situations with more modest effect sizes which are more common for complex human diseases.

While the performance of the F-statistics were similar in the current simulation study, it will be important to compare the functional F-statistics in other functional data applications. We expect our newly derived F-statistic to result in more accurate tail probabilities than the Shen and Faraway F-statistic since circumvents a degree of freedom approximation with an exact derivation. This property is important in the context of genetic association studies where small p-values are common. Further simulation studies are needed to determine if their performances are comparable in other contexts.

By applying the newly-derived functional F-statistic to neurodegenerative disease data, we were able to detect a gene-based association between HS and LATE and the *GRN* gene. While *GRN* has previously been shown to be associated with HS and, separately, other AD-related phenotypes, this is the first analysis to find a joint association between both HS and LATE and the *GRN* gene. Given that sample sizes tend be relatively small in cohorts with autopsy-derived phenotypes like NACC, the functional F-statistics should be favored for gene-based, multi-phenotype tests in these situations going forward. The original GAMuT paper compared GAMuT to a multivariate functional regression model and found GAMuT to have superior performance [31]. The approach taken to fitting the functional regression models in that study differ from ours in three important ways [40]. First, they did not use a reverse regression approach, meaning the phenotypes were modeled as a multivariate outcome. Since most clinical phenotypes tend to be correlated and typical multivariate methods assume that outcome vectors are independent of one another, we would expect a multivariate modeling approach to be sub-optimal in the presence of correlated phenotypes. Second, since a multivariate regression approach was used, there is no easy way to include mixtures of continuous and categorical phenotypes in the same analysis. Third, while the minor allele counts were smoothed before modeling, they were not flipped prior to smoothing and so there may have been some residual noise in the genotype functions thus obscuring the genomic signals. Given these deficiencies, we are not surprised that the functional regression approach performed so poorly in the context of correlated phenotypes.

The functional F-statistic approaches have several benefits over the other gene-based, multi-phenotype methods. First, since the genomic data are smoothed prior to analyzing, missing genomic data are implicitly imputed and so more genetic variants can included in the analyses. While ACAT can partially circumvent this issue by taking a complete case approach to each of the single-variant, single-phenotype analyses, GAMuT requires all missing genomic observations to be imputed prior to analyzing. Second, once the genomic data have been smoothed, the resulting genotype functions, *G*(*t*), can be evaluated at a smaller number of points to effectively reduce the dimensionality to the subsequent analyses (specifically, when the number of evaluation points is less than the number of genomic variants). Simulations showed only a marginal reduction in statistical power when the number of evaluation points was half the number of genomic variants (see Figure A.1 in the Appendices). Third, since a reverse regression approach is used for the functional F-statistics, they can easily test for associations with phenotypes of varying types (numeric, categorical, ordinal, splines, etc.). Fourth, by analyzing a gene-based, multi-phenotype association with a single regression model, no multiplicity corrections are needed for single-gene analyses. Thus, the functional F-statistic methods provide a flexible and scalable framework for conducting gene-based, multi-phenotype analyses.

In the derivation of our new F-statistic, we chose to transform the CDF values to F distributions with (*p* —*q*) and (*n* −*p*) degrees of freedom so that it would align with the F-statistic from multiple linear regression. That choice of degrees of freedom, along with the choice to transform the CDF values to F distributions, was arbitrary. While some limited simulations (not published) found that (*p* —*q*) and (*n* —*p*) degrees of freedom performed as well as, if not better than, other alternatives, there likely exists more optimal degree of freedom parameters for this F-statistic. Additionally, there may exist more optimal distributions for transforming the CDF values. Further research is needed.

While running the simulations, we came across two novel properties of ACAT [19]. First, the gene-level ACAT p-value will never exceed the lowest SNP-level p-value. So, unlike a p-value combination test like Fisher’s method which can produce a gene-level p-value that’s smaller than the minimum SNP-level p-value, ACAT acts more like a multiplicity correction in that the gene-level p-value is closer to geometric mean of the SNP-level p-values. Second, combining several ACAT p-values via ACAT without weights does not always give the same overall p-value as just combining all of the original p-values via ACAT in one pass. Notably, this only occurs when the first-level ACAT p-values consist of varying numbers of p-values and the issue can be mitigated by modifying the weights of the ACAT analyses.

In conclusion, we derived an alternative to the Shen and Faraway F-statistic for FoSR models. In the context of gene-based, multi-phenotype analyses, our newly derived functional F-statistic performed similarly to the Shen and Faraway F-statistic and both F-statistics outperformed other gene-based, multi-phenotype methods specifically in the small sample, modest effect size scenarios which are common in genetic association studies of autopsy-confirmed complex disease phenotypes like dementia. By applying the newly-derived functional F-statistic to real-world data, we were able to identify a novel association between two Alzheimer disease mimics (HS and LATE) and the *GRN* gene. Since our newly derived functional F-statistic is expected to perform better than the Shen and Faraway functional F-statistic with small p-values, it is a promising method for studies of gene-based genetic pleiotropy.

## Data Availability

1000 Genomes data are publicly available for download. National Alzheimer's Coordinating Center (NACC) and Alzheimer's Disease Genetics Consortium (ADGC) are available for research, but specific data requests must be submitted to each organization.

https://www.internationalgenome.org/data/

https://naccdata.org/

https://www.niagads.org/user/login?destination=data/request/new_request/

## Appendices

### A Varying Simulation Parameters

#### A.1 Reducing the Number of Evaluation Points for Functional Methods

**Figure A.1.**
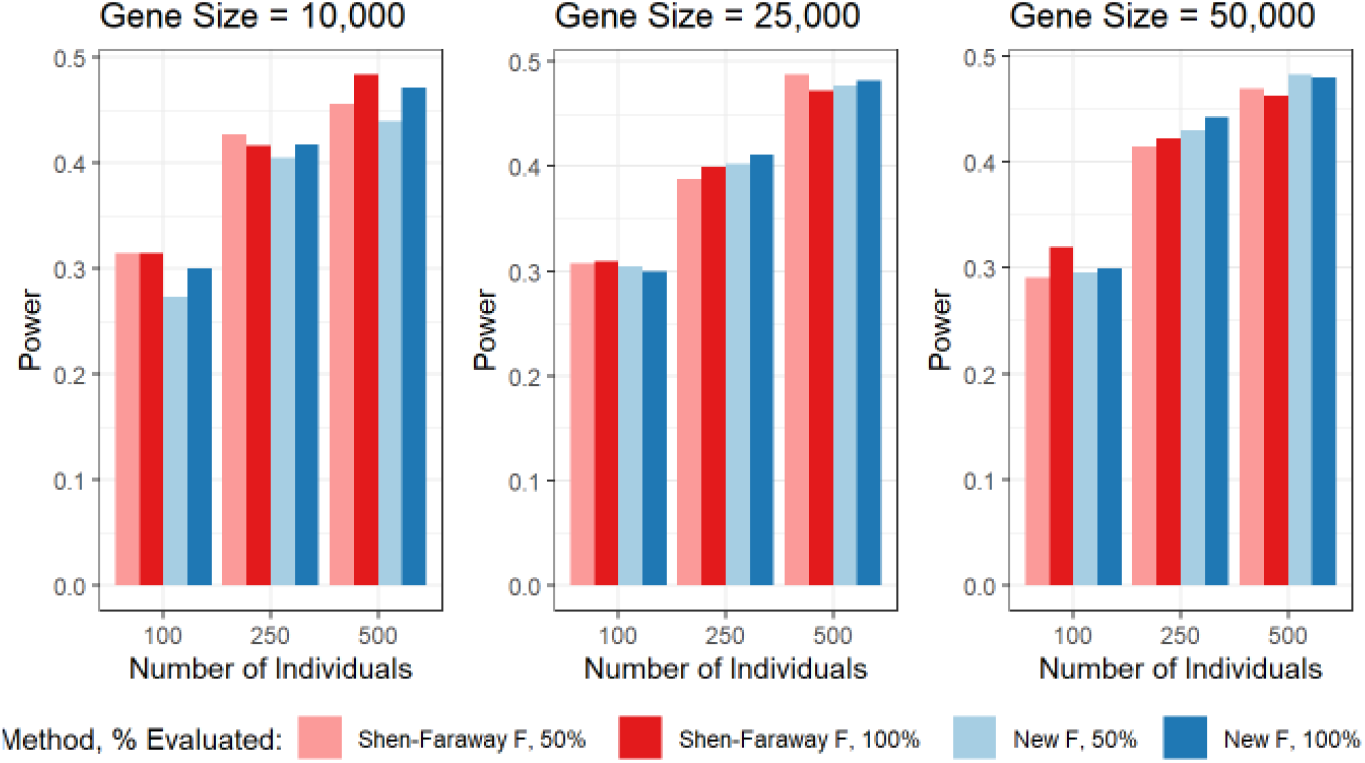
A comparison of statistical power for the functional F-statistics when the genotype functions, *G*(*t*), were evaluated at the locations of all of the included genomic variants versus half that number of equally spaced points.

## Notes

### Competing Interest Statement

The authors have declared no competing interest.

### Funding Statement

There is no external funding to report at this time.

### Author Declarations

This research is exempt.

